# Validation of questionnaires and rating scales used in medicine: Protocol for a systematic review of burnout self-reported measures

**DOI:** 10.1101/2020.06.24.20138115

**Authors:** Sandy Carla Marca, Paola Paatz, Christina Györkös, Félix Cuneo, Merete Drevvatne Bugge, Lode Godderis, Renzo Bianchi, Irina Guseva Canu

## Abstract

**Background:** In the era of evidence-based medicine, decision-making about treatment of individual patients involves conscious, specific, and reasonable use of modern, best evidences. Diagnostic tests are usually obeying to the well-established quality standards of reproducibility and validity. Conversely, it could be tedious to assess the validation studies of tests used for diagnosis of mental and behavioral disorders. This work aims at establishing a methodological reference framework for the validation process of diagnostic tools for mental disorders. We implemented this framework as part of the protocol for the systematic review of burnout self-reported measures. The objectives of this systematic review are (a) to assess the validation processes used in each of the selected burnout measures, and (b) to grade the evidence of the validity and psychometric quality of each burnout measure. The optimum goal is to select the most valid measure(s) for use in medical practice and epidemiological research.

**Methods:** The review will consist in systematic searches in MEDLINE, PsycINFO, and EMBASE databases. Two independent authors will screen the references in two phases. The first phase will be the title and abstract screening, and the second phase the full-text reading. There will be 4 inclusion criteria for the studies. Studies will have to (a) address the psychometric properties of at least one of the eight validated burnout measures (b) in their original language (c) with sample(s) of working adults (18 to 65 years old) (d) greater than 100. We will assess the risk of bias of each study using the Consensus-based Standards for the selection of health Measurement Instruments checklist. The outcomes of interest will be the face validity, response validity, internal structure validity, convergent validity, discriminant validity, predictive validity, internal consistency, test-retest reliability, and alternate form reliability, enabling assessing the psychometric properties used to validate the eight concerned burnout measures. We will examine the outcomes using the reference framework for validating measures of mental disorders. Results will be synthetized descriptively and, if there is enough homogenous data, using a meta-analysis.

**Ethics and dissemination:** We will publish this review in a peer-reviewed journal. A report will be prepared for the health practitioners and scientists and disseminated through the Network on the Coordination and Harmonization of European Occupational Cohorts (https://www.cost.eu/actions/CA16216, http://omeganetcohorts.eu/) and the Network of scientists from Swiss universities working in different areas of stress (https://www.stressnetwork.ch/).

**PROSPERO registration number:** CRD42019124621

## BACKGROUND

### Rationale

In the era of evidence-based medicine (EBM), decision-making about treatment of individual patients involves conscious, specific, and reasonable use of modern, best evidences (1). The purpose of EBM is ultimately to provide patients with the best treatment solutions. Thus, EBM helps avoid mistakes in the course of treatment and raises the quality and the cost-effectiveness of health care. Diagnosis and prognosis, two basic aspects of medicine and paramedicine, provide valuable information enabling patients and professionals to make decision. The results of diagnostic and prognostic processes must be as correct as possible, as they can have far-reaching consequences. The application of the EBM methods in diagnostic and prognostic processes used in healthcare is thus essential (2).

EBM requires from the physician the ability to search the medical literature and the skills in the interpretation of epidemiological and statistical results. However, evaluating the quality of a given study can be challenging in some cases, depending on the nature of the diagnostic test, the study design and statistics used. For instance, diagnostic tests involving measurable functional, biological or morphological changes of clinical significance usually obey to well established quality standards of reproducibility and validity and are relatively easy to compare based on their predictive values, sensitivity and specificity (3). In contrast, validity studies of tests in questionnaire format, commonly used for the diagnosis of mental and behavioral disorders, are more challenging to assess. Diagnostic questionnaire assessing mental disorders should obey to a number of methodological standards, such as psychometric properties, as part of its validation process (4). However, terms that denominate the psychometric properties have rather broad, sometimes vague definitions, while the statistical methods for their assessment vary widely across publications (4-11). Moreover, available methodological guidelines are heterogeneous and generally incomplete. Some of them are even contradictory (4, 6, 7). To date, no consensual methodological guideline exists for the whole validation process of mental health questionnaires and rating scales used for screening and diagnosis of mental disorders. The currently available standards focus on the methodological quality of single studies reporting diagnostic accuracy and psychometric properties. Examples of those standards are the Quality Assessment of Diagnostic Accuracy Studies (QUADAS) (12) or the Standards Reporting of Diagnostic Accuracy Studies (STARD) (13). The Consensus-based Standards for the selection of health Measurement Instruments (COSMIN) (14) is often used for the qualitative evidence appraisal in the systematic reviews. However, the latest is rather unhelpful from the statistical point of view.

This lack of harmonization regarding acceptable validity standards or criteria for various mental health questionnaires directly challenges the EBM application in diagnosis and subsequently in treatment of mental disorders, in particular, among non-specialized health professionals. In order to remedy this situation, we have established a general reference framework for the validation process of diagnostic tools for mental disorders, including self-reported measures of burnout. The burnout syndrome remains ill-defined and nosologically uncharacterized (15). Despite its increasing importance (16), burnout syndrome still has no consensual definition, which makes it difficult to manage. Maslach and Jackson (17) proposed the most prominent definition of burnout: a psychological syndrome that occurs in professionals who work with other people in challenging situations that is measured through three domains: 1-emotional exhaustion 2-depersonalisation and 3-personal accomplishment. From this definition, Maslach developed a first burnout measure: the Maslach Burnout Inventory (MBI). Apart from the MBI, a meta-analysis by O’Connor et al. (18) cited six other validated burnout measures: the Pines Burnout Measure (BM), the OLdenburg Burnout Inventory (OLBI), the Copenhagen Burnout Inventory (CBI), the Professional Quality of Life Scale (ProQOL III), the Psychologists Burnout Inventory (PBI), the Children’s Services Survey (CSS), and the Organizational Social Context Scale (OCS). Considering that psychological syndromes measures are heterogeneous, a closer look to the validation process of the currently used burnout measures should give insight on their legitimacy in medical practice and research.

### Objectives

This article aims at presenting our methodological reference framework for the validation process of diagnostic tools for mental disorders as part of the protocol for our systematic review of burnout self-reported measures. The objectives of this systematic review are to assess the validation processes used in each of the selected burnout measures and to grade the evidence of the validity and psychometric quality of each burnout measure to select the most valid one(s) for use in medical practice and epidemiological research.

## METHODS AND ANALYSIS

We developed the protocol according to the Preferred Reporting Items for Systematic Reviews and Meta-Analyses (PRISMA) recommendations. We registered the protocol with the International Prospective Register of Systematic reviews (registration number CRD42019124621).

### Reference framework for the validation process of diagnostic tools for mental disorders

This framework is provided in Supplementary material Table 1, organized in four columns, as follows: 1-psychometric validity criteria, 2-their definitions, 3-the methods commonly used to analyze them, and 4-the resulting statistical estimates and indices as well as the objective criteria for their respective interpretation. To construct this framework, we completed the demarche initiated by the French National Institute of Research on Security (INRS) for a comparative analysis of different scales and tools used for assessing psychosocial risks available in French language (4). First, we listed as exhaustively as possible the psychometric validity criteria and their definitions, using handbooks and published guidelines (4-11, 14, 15, 17-44). Second, we sorted the validity criteria, according to their most consensual denomination and definition and grouped them by sub-types according to Bolarinwa (6). Third, we filled the third and fourth columns of the table with appropriate analyses and indices’ interpretation for each validity criterion, using handbooks and published methodological guidelines (4-6, 8-11, 19-33, 35-44). Forth, we submitted the completed table of our framework to two independent experts with strong psychometric skills for critical review of the retained definitions, the completeness of the methods, and the appropriate choice of interpretation criteria. Finally, after discussion of the reviewers’ comments and getting consensus, we produced a current version of the framework. We consider it as a methodological referential because it allows non-specialized health professionals and researchers to understand and to correctly interpret the overall and specific validity criteria of a diagnostic tool for mental disorders, whatever the study design and statistical method used for its validation. Thanks to its multiple entries, it is possible to shift through validation studies by picking up terms about either validity criteria (20 criteria), analytical methods (21 methods) or the resulting indices and statistics grouped into 19 categories. Because of its analytical exhaustiveness and completeness for the three elements of the validation of diagnostic tests (i.e., validity, reproducibility and sensitivity), it constitutes a useful framework for quality appraisal of diagnostic tests for mental disorders.

**Table 1.**
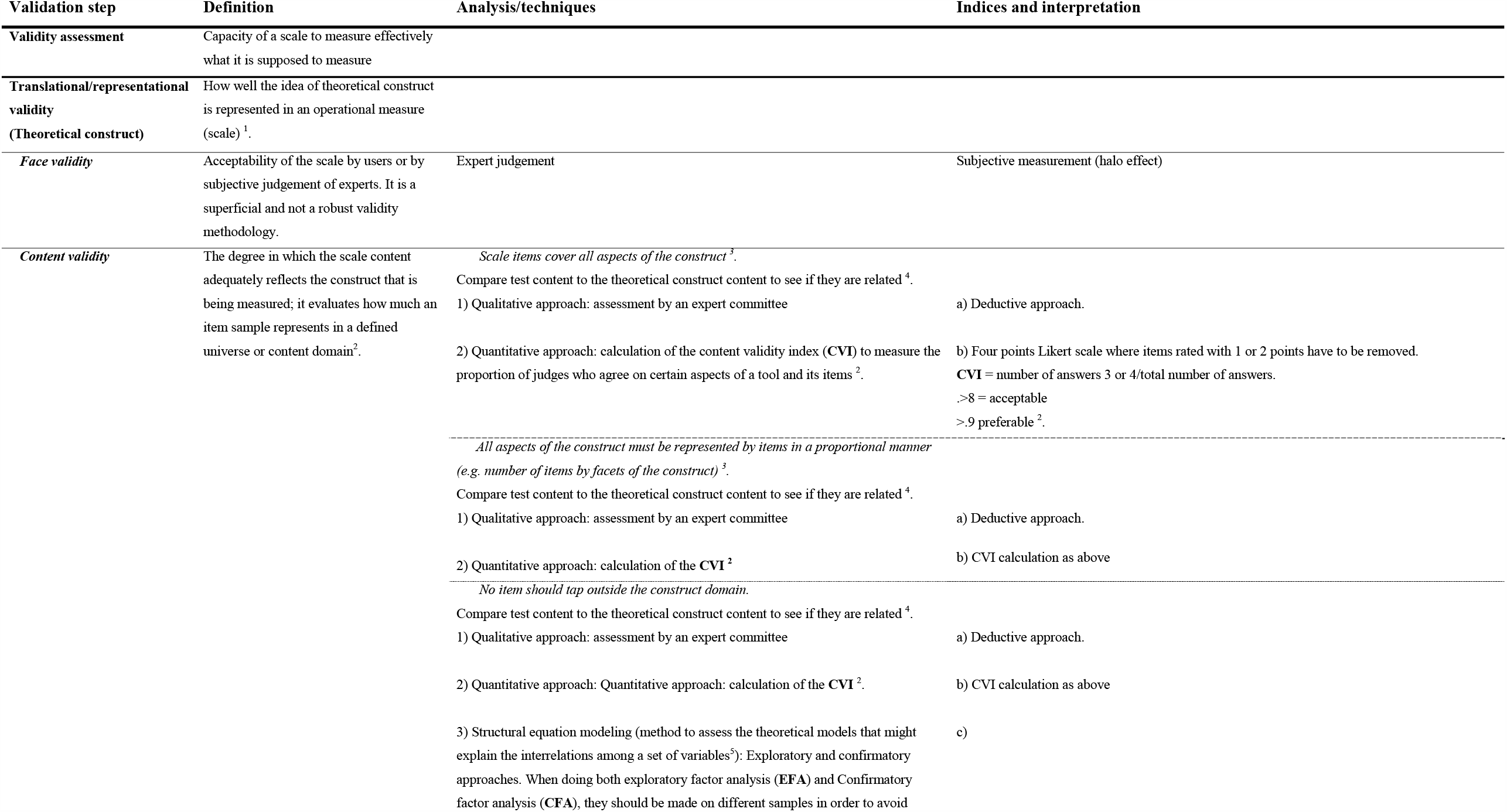
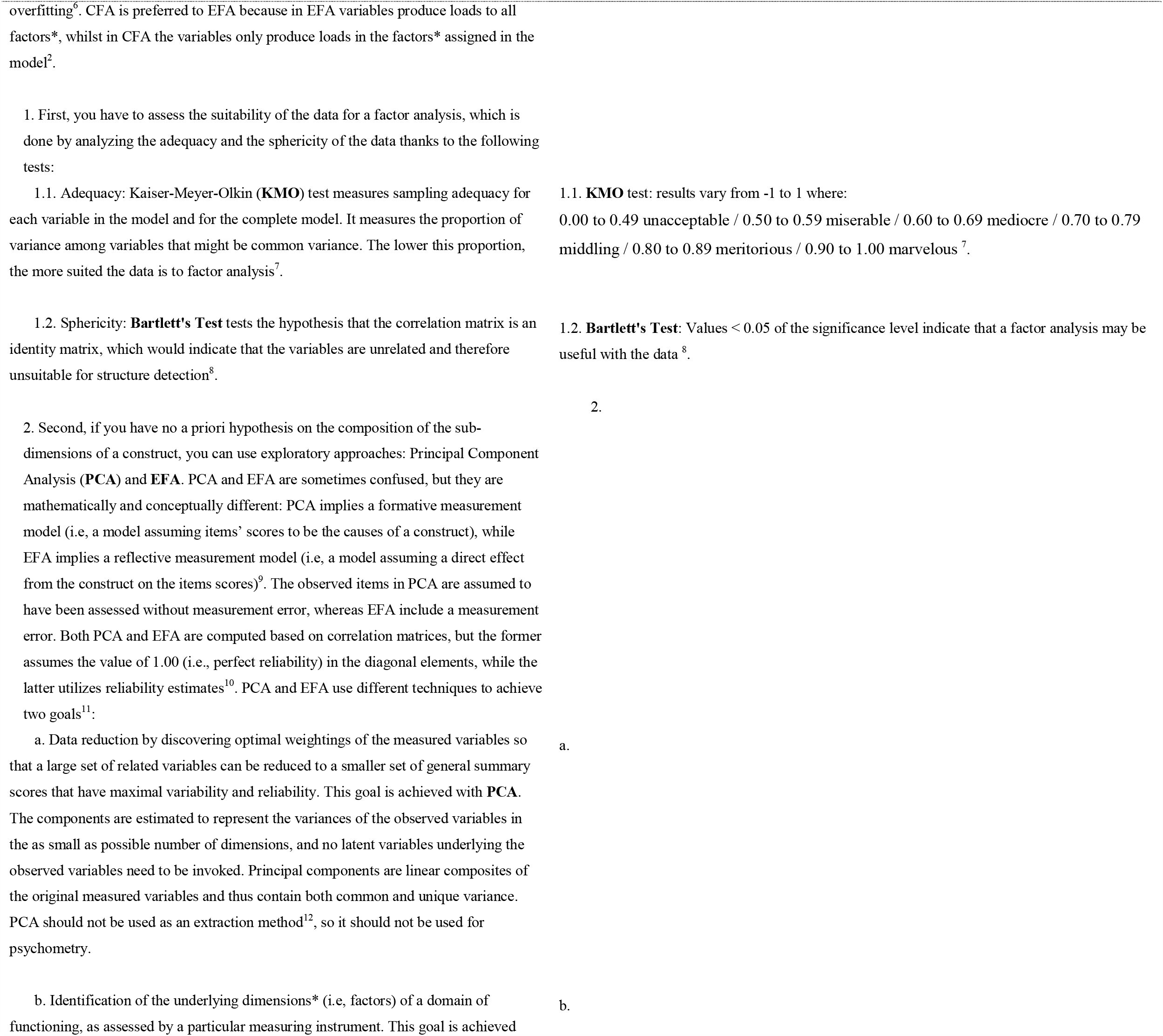
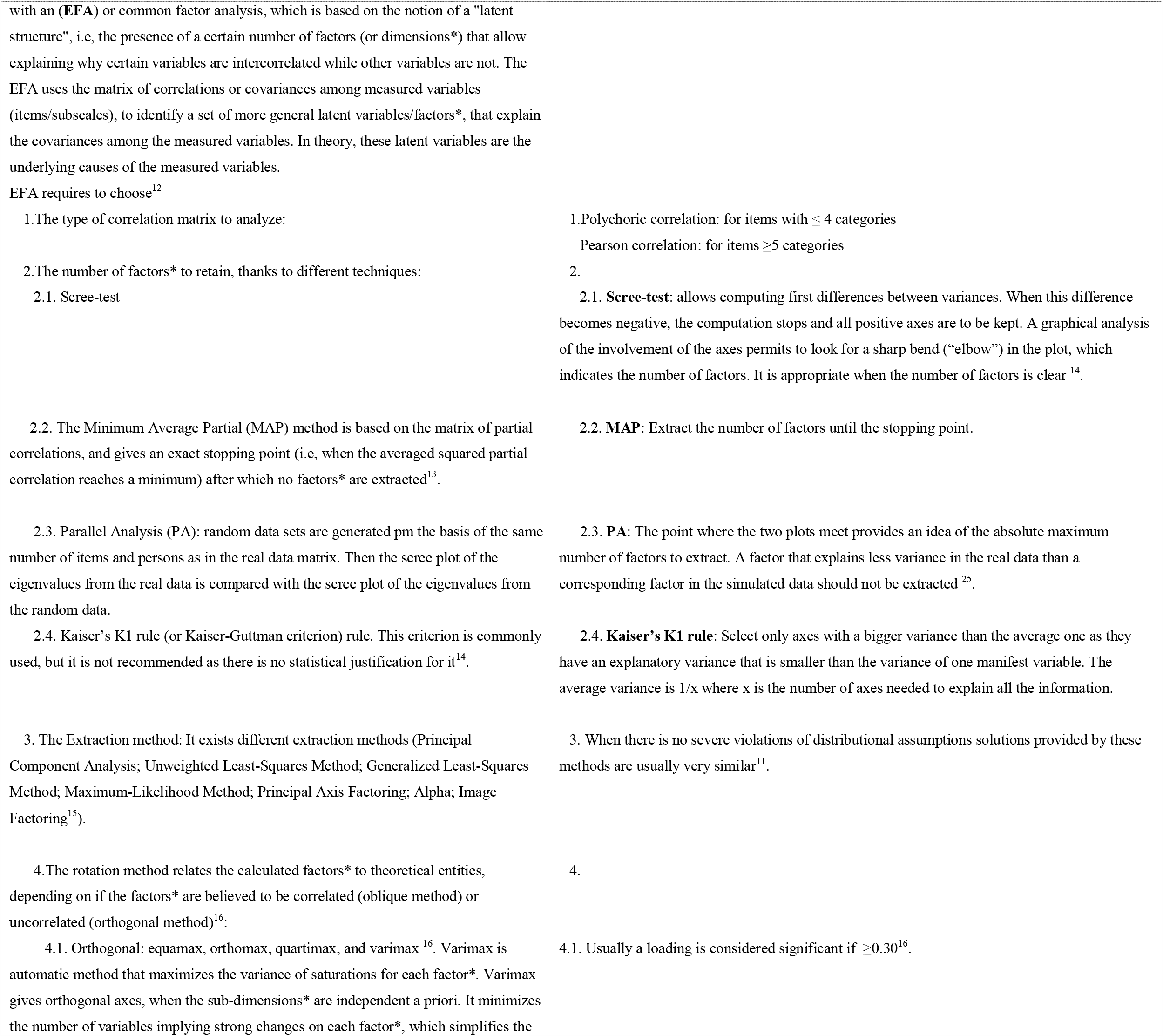
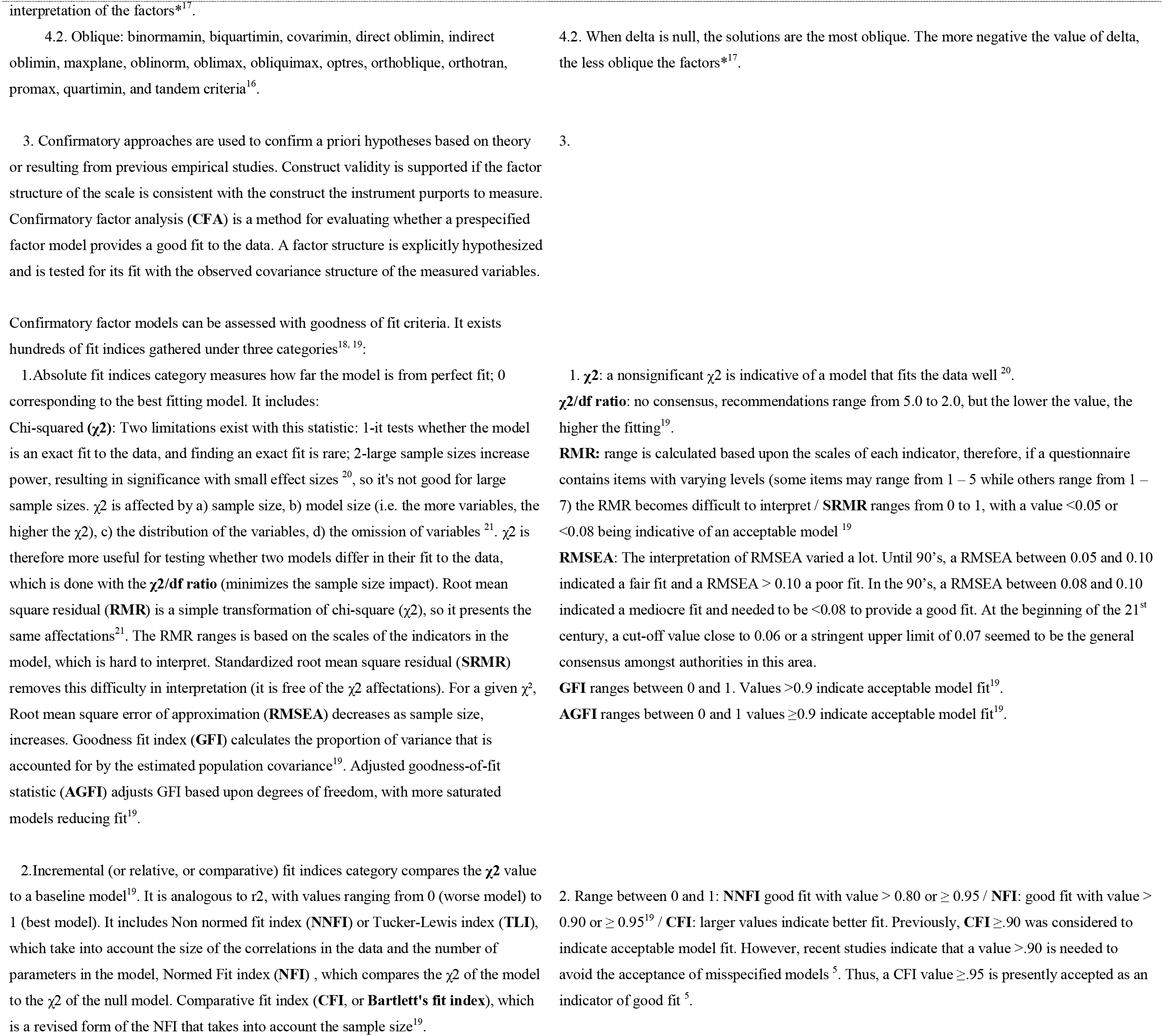
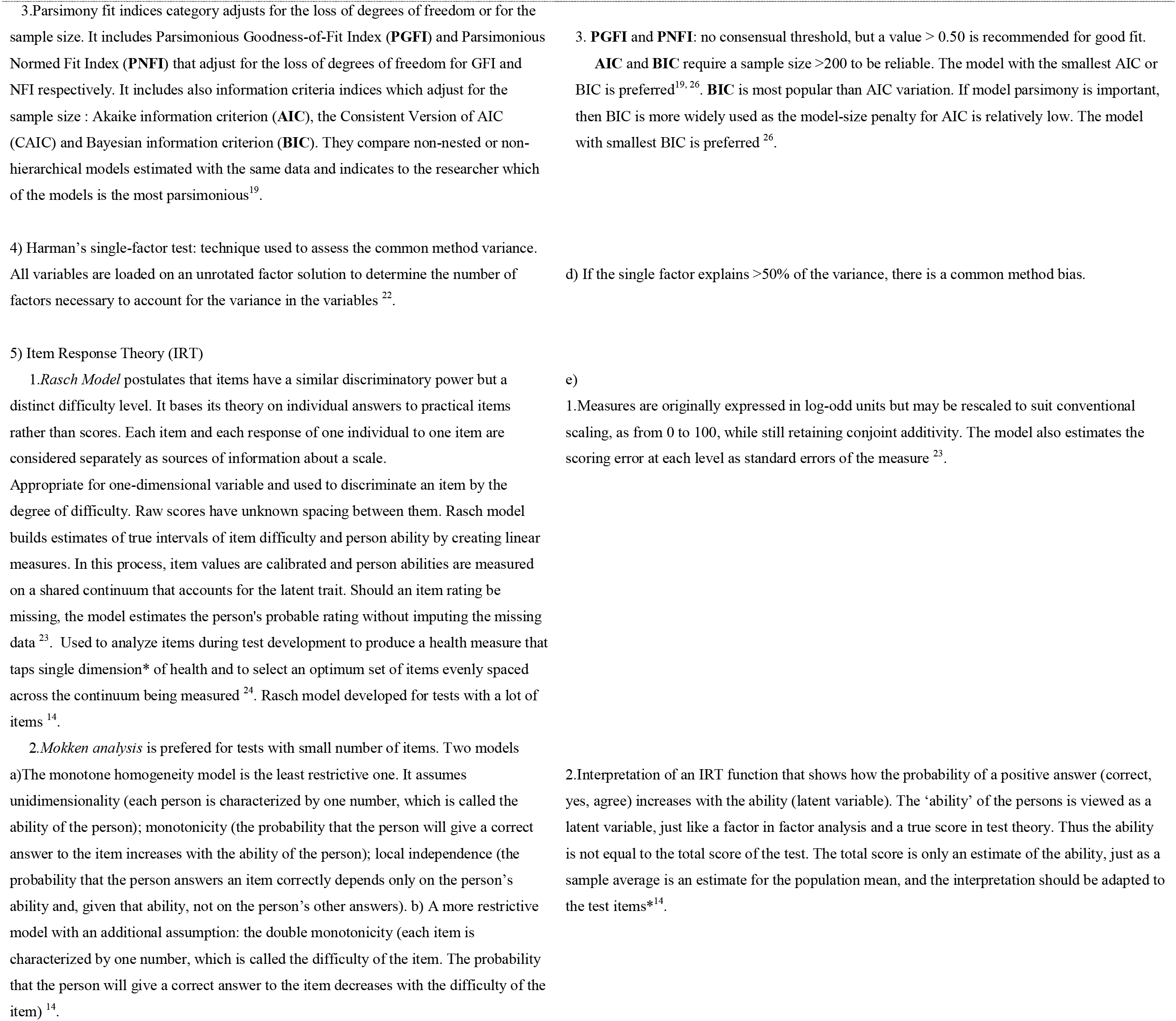
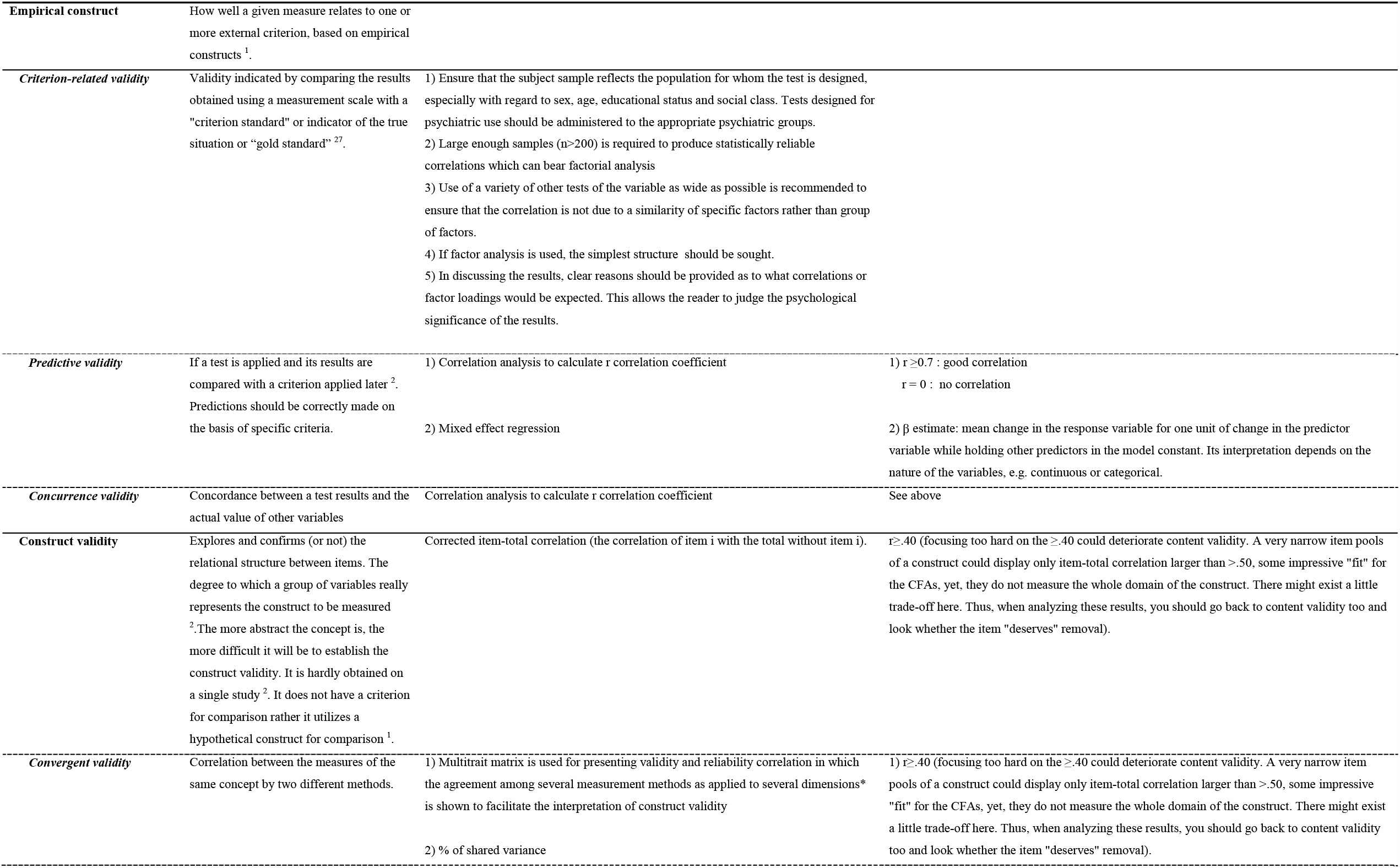
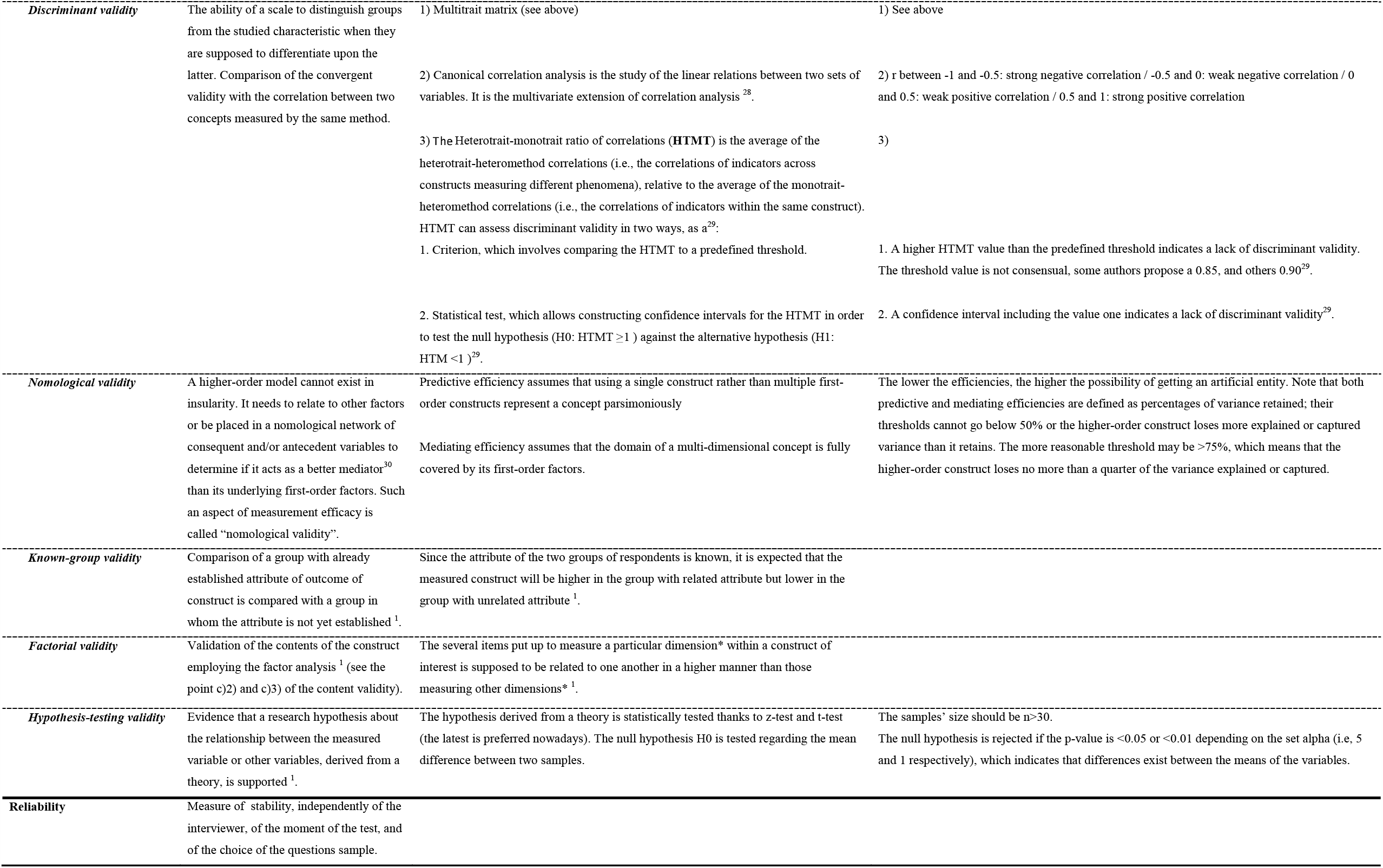
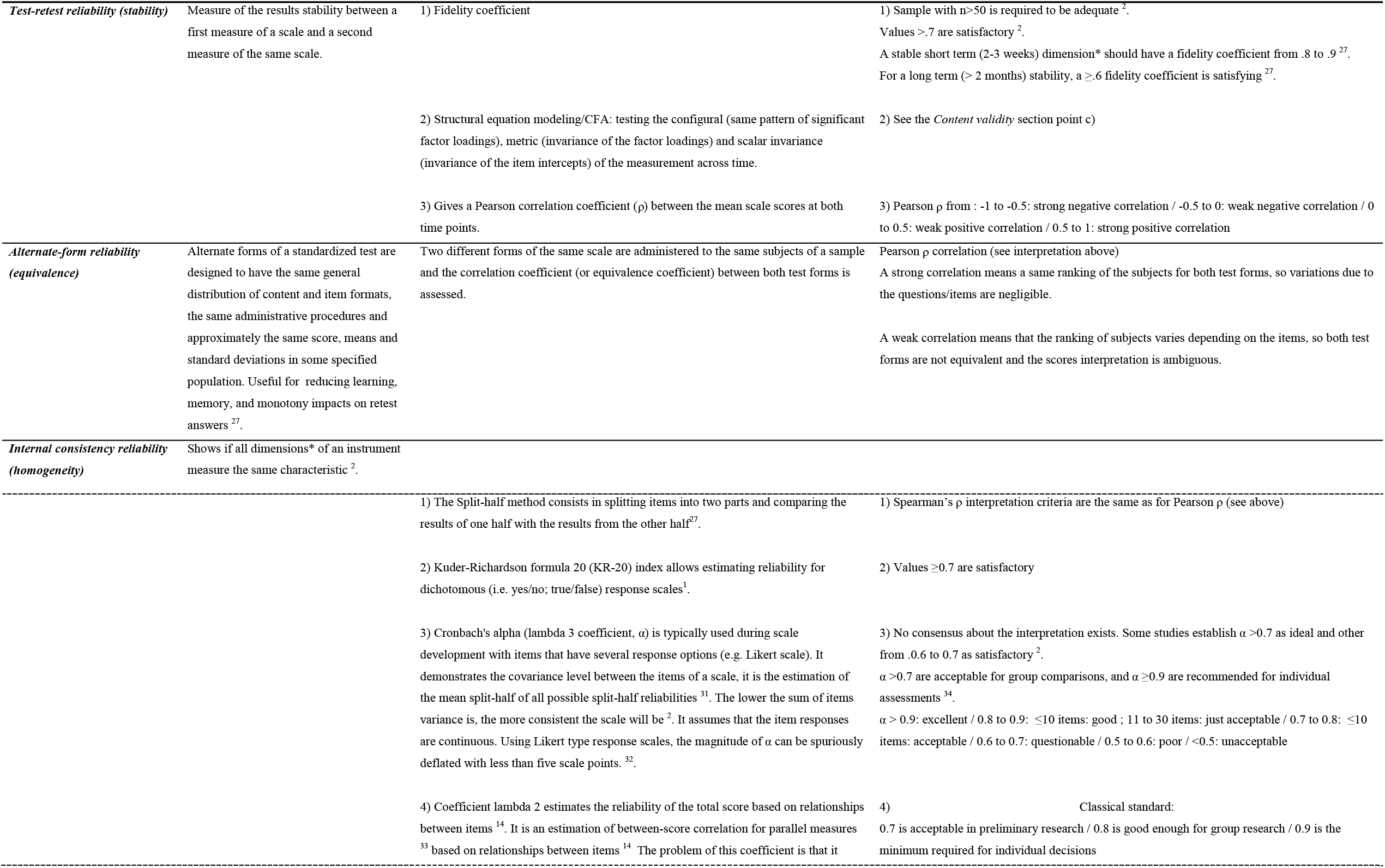
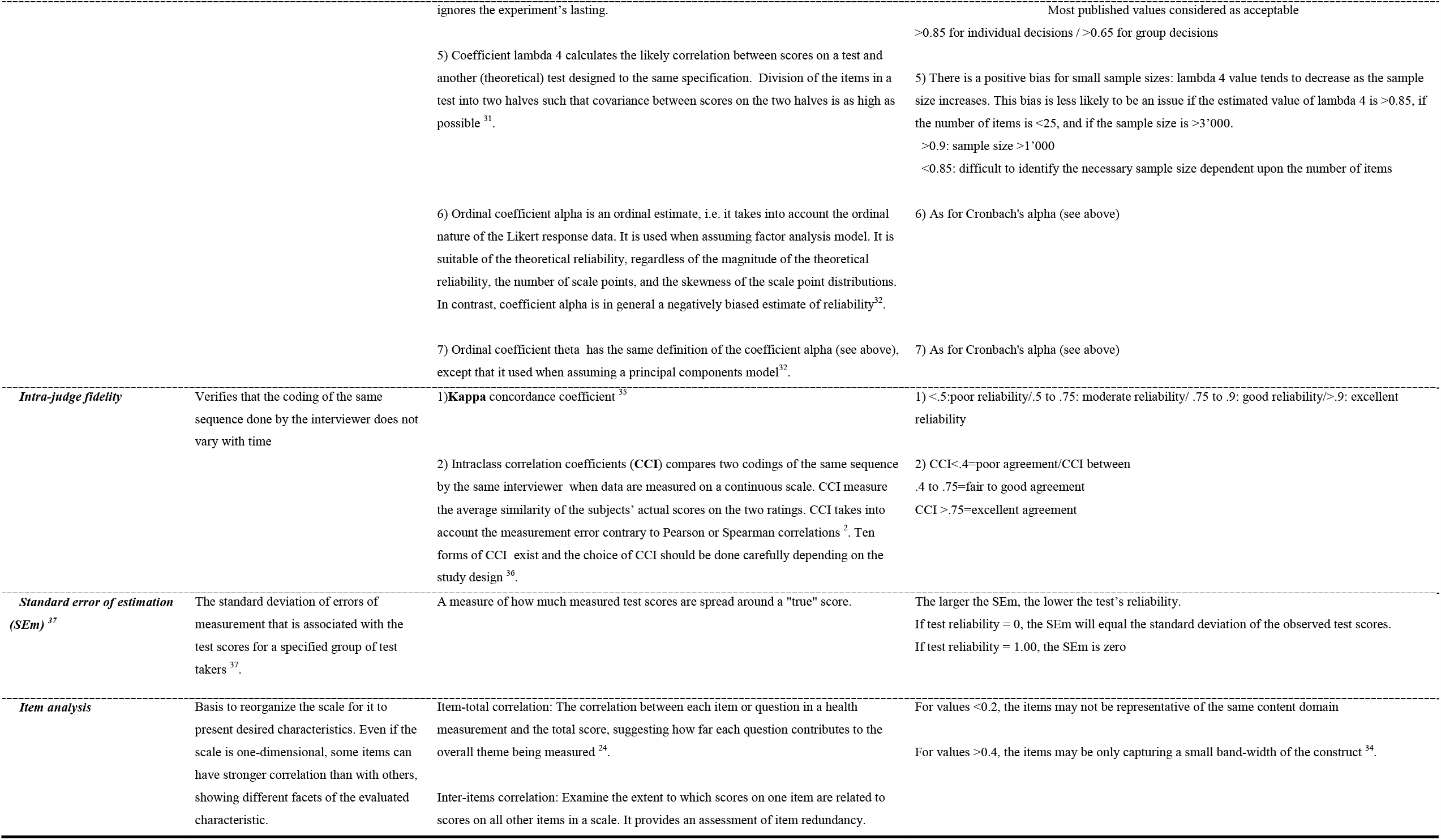
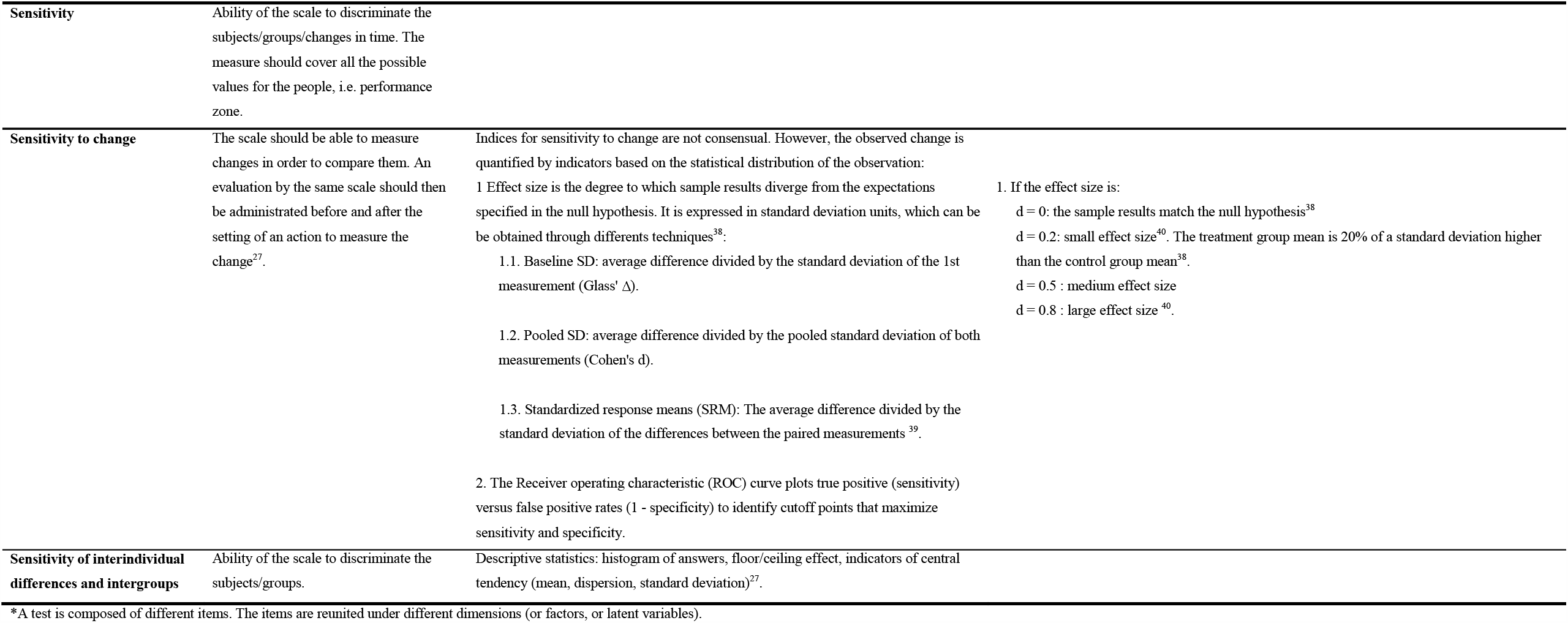
Referential framework for validation of questionnaires and rating scales

### Eligibility criteria

We will include 1-studies with quantitative methodology; 2-published in the original scientific article formats; 3-adressing the psychometric properties of at least one above-mentioned burnout measures in its original (not translated) version; 4-with sample size of at least of 100 participants. We will exclude 1-studies that do not meet the inclusion criteria; 2-studies for which no abstract and full text could be found; 3-studies where one of the eight burnout measures was used as a reference against another one, not included in this review; 4-studies where a translated version of burnout measure was used (e.g., translational validity and cross-cultural studies); 5-studies in which quantitative data on reliability or validity were missed; 6-studies where participants were not professionally employed (e.g., students, medical residents).

#### Participants

We will include studies with working adult participants aged between 18 and 65 years old. We will exclude studies where participants had no professional occupation (e.g., students, medical residents).

#### Exposures/Interventions

This review is focused on the psychometric properties and validity of the selected burnout self-reported measures. It would not consider the exposures or predictors of burnout in workers.

#### Comparators

We will consider measures of depression, anxiety, and somatic disorders as comparators to assess the discriminant validity of burnout measures.

#### Outcome measures

The outcome are the psychometric properties used to validate the eight aforementioned burnout measures: Face validity; Response validity; Internal structure validity; Convergent validity; Discriminant validity; Predictive validity; Internal consistency reliability; Test-retest reliability; Alternate form reliability.

#### Time frame

As we include quantitative studies reporting one of the above-mentioned outcomes, we expect different time frames to be used in the selected studies. Thus, no restriction to any particular time frame will be applied.

#### Setting

Given that the study population consists of working adults, all occupational settings will be considered. If enough homogenous data are available per type of occupation, we will perform additional analysis for specific occupational settings (e.g., health care, education).

#### Language

There will be no language restriction

#### Information sources

Systematic literature search will be performed for the period from 1980 to 2018 (September). This period was determined with the argument that the first validated measure of burnout was published in 1981 with the MBI (17). We will use three databases to search for studies of interest via the online catalog of databases OVID interface: the Medical Literature Analysis and Retrieval System Online (MEDLINE) database, the world-class resource for abstracts and citations of behavioral and social science research PsycINFO database, and the Excerpta Medica database (EMBASE). In addition, we will check the reference lists from articles and reviews retrieved in our electronic search for any additional studies to include.

### Search strategy

An experienced librarian will review the search strategy. It will consist of free-text words to specify three search strings: terms focusing on the burnout measure of interest (e.g., MBI), terms related to the validation of the measure, and a combination of the two first search strings results. Finally, one additional search string will consist of removing duplicates.

### Study records

#### Data management

We will import the collected studies in the bibliography software EndNote X8.

#### Selection process

Two independent reviewers will screen the references to eliminate the eventual remaining duplicates within each database. They will also eliminate duplicates between databases. They will screen the remaining articles based on their title and abstract. They will retain or reject the articles based on the above-mentioned inclusion and exclusion criteria. The two reviewers will then screen the remaining articles based on full-text reading. They will discuss any discrepancies and if needed, ask a third reviewer to arbitrate the decision. A reviewer will illustrate the selection process with a flowchart following the PRISMA guidelines.

#### Data collection process

To elaborate a standardized data extraction form convenient for all kinds of study design and methods applied; we will use our reference framework for the validation process of diagnostic tools for mental disorders (Table 1). Each burnout measure will have its own exemplary of data extraction form (MS Excel file) that will be filled with studies’ data concerning the burnout measure in question. Two independent reviewers will test the form using articles on different burnout measures. They will discuss any discrepancies and if needed, they will ask a third reviewer to arbitrate the decision and add clarification. This process will continue until complete agreement is reached between both reviewers on the finalized data extraction form. The data of the included studies will be extracted by one of two reviewers. A second reviewer will crosscheck a random 20% sample of the extracted data. The missing data will be identified by a code depending on the reason why they are missing (e.g., not assessed, not reported). The data extraction process will provide additional validation of the referential framework completeness.

### Data items

The extracted data will concern studies’ identification (i.e., authors, year of publication, journal, and title); samples’ characteristics (i.e., size, gender ratio, age, occupational activity, participation rate, representativity, burnout scores’ distribution); burnout measures’ characteristics (i.e., name, version, number of items, number of domains, domains’ names); and statistical methods used for assessing the psychometric properties outcome.

### Outcomes and prioritization

The outcomes of interest will be the face validity, response validity, internal structure validity, convergent validity, discriminant validity, predictive validity, internal consistency, test-retest reliability, and alternate form reliability. Those criteria will enable to assess the psychometric properties used to validate the eight concerned burnout measures.

### Risk of bias in individual studies

Two reviewers will independently assess the quality of each study using the COSMIN checklist (14). They will discuss any discrepancies, and they will resort to the arbitration of a third reviewer if needed.

### Data synthesis

#### Descriptive analyses

We will interpret the quantitative based on our methodological reference framework. We will create a narrative synthesis of the findings from the included studies. We will structure this synthesis around the burnout measure, the target population characteristics, and the type of outcome.

We plan to carry out subgroup analysis on the primary outcomes by grouping studies based on the following: 1-Burnout self-reporting measure: MBI, BM, OLBI, CBI, ProQOL III, PBI, CSS, and OCS.

2-Burnout domain: Emotional exhaustion, Depersonalization, Personal accomplishment (MBI); Physical exhaustion, Mental exhaustion (BM); Disengagement, Exhaustion (cognitive and physical) (OLBI); Professional exhaustion, Personal Exhaustion, Relational Exhaustion (CBI); Compassion fatigue burnout (ProQOL III); Aspects of control, Support in the work setting, Type of negative clientele, Overinvolvement with the client (PBI); Emotional exhaustion (CSS); Culture, Climate, Work attitudes (OCS).

3-Participants’ characteristics: gender, age, and burnout score.

#### Meta-analyses

There might be a limited scope for meta-analysis. There will be a range of different factors and outcomes measured and reported across existing studies. However, we will pool summary estimates in form of multiple logistic regression coefficients whenever possible. We will do it for study overlapping in terms of outcome measures, for at least one of the burnout domains. Since the participants in the various studies might be construed as coming from the same population (workers) or from different populations (i.e., according to each study’s inclusion criteria) we will use a fixed effects model.

#### Meta-biases

According to standard practice in meta-analysis, the first step will be to represent the data as forest plots including the I-square that estimates the percentage of the between-study heterogeneity. If the latter is very large, this means that the between-study heterogeneity is much larger than the between-subject heterogeneity and any attempt of obtaining a reference value for individual subjects will not be valid(45).

##### Assessment of publication bias

We will produce funnel plots to investigate possible publication bias, as recommended in the epidemiological literature.

##### Assessment of heterogeneity

For each model, heterogeneity will be assessed by quantifying the inconsistency across studies using I^2^ statistic greater than 50% as criterion. If heterogeneity is identified, potential causes will be explored (e.g. clinical and/or methodological diversity). We will try to clarify heterogeneity via subgroup analysis, but if it cannot be explained (i.e. there is considerable variation in the results), then a meta-analysis using a random-effect model will be conducted. We will exclude studies with a high risk of bias to determine the extent to which the synthesized results are sensitive to risk of bias. Statistical analysis will be performed using STATA software, 16th version.

#### Confidence in cumulative evidence

The strength of the evidence for the relationship between different risk factors and burnout onset will be assessed using the Grading of Recommendations Assessment, Development and Evaluation (GRADE) approach. It will allow to rate the certainty of a body of evidence as suggested by GRADE guidelines 18 (46). We will use a checklist designed by Meader et al. (2014) (47) to improve consistency and reproducibility of our GRADE assessment. The results will be presented using the GRADE Summary of Findings Tables and Evidence Profiles (48).

## Data Availability

All data ate provided in the Table 1

## Acknowledgements

The authors thank Aline Sager, the Unisanté/DSTE librarian.

## Notes

**Funding** University of Lausanne and University of Bern BNF – National Qualification Program funded the salary of young researchers (PP and SCM); European Cooperation in Science & Technology (COST Action CA16216), OMEGA-NET: Network on the Coordination and Harmonization of European Occupational Cohorts covered the meetings and travel expenses as well as the open access publication costs.

### Competing Interest Statement

The authors have declared no competing interest.

### Clinical Protocols

https://www.crd.york.ac.uk/prospero/display_record.php?RecordID=124621

### Funding Statement

University of Lausanne and University of Bern National Qualification Program funded the salary of young researchers (PP and CG); European Cooperation in Science & Technology (COST Action CA16216), OMEGA-NET, Network on the Coordination and Harmonization of European Occupational Cohorts covered the meetings and travel expenses as well as the open access publication costs.

### Author Declarations

As it is a systematic review protocol and methodology for validity assessement of pation-reported outcome measures, no approval from ethics commettey was necessary

## REFERENCES

1. Sackett DL, Rosenberg WMC, Gray JAM, Haynes RB, Richardson WS. Evidence-based medicine: what it is and what it isn’t. BMJ. 1996;312:71–2.

2. Newman TB, Kohn MA. Evidence-Based Diagnosis: Cambridge: Cambridge University Press; 2009.

3. Bouter LM, Zielhuis GA, Zeegers MP. Textbook of Epidemiology: Bohn Stafleu van Loghum; 2018. 228 p.

4. Langevin V, François M, Boini S, Riou A. Les questionnaires dans la démarche de prévention du stress au travail.. Documents pour le Médecin du Travail. 2011;125(1er trimestre 2011):23–35.

5. André N, Loye N, Laurencelle L. La validité psychométrique: un regard global sur le concept centenaire, sa genèse, ses avatars. Mesure et évaluation en éducation. 2015;37(3).

6. Bolarinwa OA. Principles and methods of validity and reliability testing of questionnaires used in social and health science researches. Niger Postgrad Med J. 2015;22(4):195–201.

7. Geisinger KF, Bracken BA. APA Handbook of Testing and Assessment in Psychology: American Psychological Association; 2013.

8. McDowell I. Measuring Health: A guide to rating scales and questionnaires Oxford University Press; 2006.

9. Nunally JC. Psychometric Theory. 2nd ed. New York: McGraw-Hill; 1978. 701 p. 10.

10. Bernaud J-L. Introduction à la psychométrie. Paris: France: Dunod; 2007.

11. Souza AC, Alexandre NMC, Guirardello EB. Psychometric properties in instruments evaluation of reliability and validity. Epidemiol Serv Saude. 2017;26(3):649–59.

12. Wade R, Corbett M, Eastwood A. Quality assessment of comparative diagnostic accuracy studies: our experience using a modified version of the QUADAS-2 tool. Res Synth Methods. 2013;4(3):280–6.

13. Cohen JF, Korevaar DA, Altman DG, Bruns DE, Gatsonis CA, Hooft L, et al. STARD 2015 guidelines for reporting diagnostic accuracy studies: explanation and elaboration. BMJ Open. 2016;6(11):e012799.

14. Terwee CB, Prinsen CA, Chiarotto A, Westerman MJ, Patrick DL, Alonso J, et al. COSMIN methodology for evaluating the content validity of Patient-Reported Outcome Measures: a Delphi study. Quality Of Life Research. 2018;27(5):1159–70.

15. Guseva-Canu I, Mesot O, Györkös C, Mediouni Z, Mehlum IS, Bugge MD. Burnout syndrome in Europe: towards a harmonized approach in occupational health practice and research. Industrial Health. 2019 (in press).

16. Shanafelt TD, Dyrbye LN, West CP. Addressing Physician Burnout: The Way Forward. JAMA. 2017;317(9):901–2.

17. Masclach C, Jackson SE. The measurement of experienced burnout. Journal of Occupational Behaviour. 1981;2(99):99–113.

18. O’Connor K, Muller ND, Pitman S. Burnout in mental health professionals: A systematic review and meta-analysis of prevalence and determinants. Eur Psychiatry. 2018;53:74–99.

19. Cameron AC, Trivedi PK. Microeconometrics: Methods and applications. New York: Cambridge University Press; 2005. 1034 p.

20. IBM Knowledge Center. KMO and Bartlett’s Test [Available from: https://www.ibm.com/support/knowledgecenter/en/SSLVMB_24.0.0/spss/tutorials/fac_telco_kmo_01.html.

21. IBM Knowledge Center. Rotation d’analyse factorielle [Available from: https://www.ibm.com/support/knowledgecenter/fr/SSLVMB_23.0.0/spss/base/idh_fact_rot.html.

22. Granger CV. Rasch Analysis is Important to Understand and Use for Measurement Buffalo 2008 [Available from: https://www.rasch.org/rmt/rmt213d.htm.

23. Kenny DA. Measuring Model Fit 2015 [Available from: http://davidakenny.net/cm/fit.htm.

24. Hooper D, Coughlan J, Mullen MR. Structural equation modelling; Guidelines for determining model fit. J Res Natl Inst Stand Technol. 2008;6(1):53–60.

25. Hu LT, Bentler PM. Cutoff criteria for fit indexes in covariance structure analysis: Conventional criteria versus new alternatives. Structural Equation Modeling: A Multidisciplinary Journal. 1999;6(1):1–55.

26. Brown JD. Choosing the right type of rotation in PCA and EFA. JALT Testing & Evaluation SIG Newsletter. 2009;13(3):20-5. 27.

27. Mandrekar JN. Measures of interrater agreement. Journal of Thoracic Oncology. 2011;6(1):6–7.

28. Newsom JT. Some clarification and recommendations on fit indices. 2018.

29. Koo TK, Li MY. A Guideline of Selecting and Reporting Intraclass Correlation Coefficients for Reliability Research. J Chiropr Med. 2016;15(2):155–63.

30. Becker LA. Effect Size (ES). 2000.

31. Liu L, Li C, Zhu D. A new approach to testing nomological validity and its application to a second-order measurement model of trust. Journal of the Association for Information Systems. 2012;13(12):950–75.

32. Michalos AC. Encyclopedia of Quality of Life and Well-Being Research. Prince George, BC, Canada: SpringerReference; 2014. 7347 p.

33. Newton PE, Shaw SD. Validity in Educational & Psychological Assessment. London: SAGE; 2014. 280 p.

34. Ray SL, Wong C, White D, Heaslip K. Compassion satisfaction, compassion fatigue, work life conditions, and burnout among frontline mental health care professionals. Traumatology. 2013;19(4):255–67.

35. Arcanger S. Analyse de Données: TP3. 2008-2009.

36. MEDALC easy-to-use statistical software. Responsiveness. Unkown [Available from: https://www.medcalc.org/manual/responsiveness.php.

37. NCSS Statistical Software. Canonical correlation. Unkown [Available from: https://ncss-wpengine.netdna-ssl.com/wp-content/themes/ncss/pdf/Procedures/NCSS/Canonical_Correlation.pdf.

38. Université de Toulouse. Analyse en Composantes Principales (ACP). Unkwown.

39. Statistics How To: Statistics for the rest of us! Kaiser-Meyer-Olkin (KMO) Test for Sampling. 2016 [Available from: https://www.statisticshowto.datasciencecentral.com/kaiser-meyer-olkin/.

40. Statistics How To: Statistics for the rest of us! Standard Error of Measurement (SEm): Definition, Meaning. 2016 [Available from: https://www.statisticshowto.datasciencecentral.com/standard-error-of-measurement/.

41. Vacha-Haase T, Thompson B. How to estimate and interpret various effect sizes. Journal of Counseling Psychology. 2004;51(4):473–81.

42. Weston R, Gore PA. A Brief Guide to Structural Equation Modeling. The Counseling Psychologist. 2016;34(5):719–51.

43. Wikipedia. Confirmatory factor analysis - Goodness of fit index and adjusted goodness of fit index. [Available from: https://en.wikipedia.org/wiki/Confirmatory_factor_analysis#Goodness_of_fit_index_and_adjusted_goodness_of_fit_index.

44. Zumbo BD, Gadermann AM, Zeisser C. Ordinal Versions of Coefficients Alpha and Theta for Likert Rating Scales. Journal of Modern Applied Statistical Methods. 2007;6(1):21–9.

45. Higgins JP, Altman DG, Gøtzsche PC, Jüni P, Moher D, Oxman AD, et al. The Cochrane Collaboration’s tool for assessing risk of bias in randomised trials. Bmj. 2011;343:d5928.

46. Schunemann HJ, Cuello C, Akl EA, Mustafa RA, Meerpohl JJ, Thayer K, et al. GRADE guidelines: 18. How ROBINS-I and other tools to assess risk of bias in nonrandomized studies should be used to rate the certainty of a body of evidence. J Clin Epidemiol. 2018.

47. Meader N, King K, Llewellyn A, Norman G, Brown J, Rodgers M, et al. A checklist designed to aid consistency and reproducibility of GRADE assessments: development and pilot validation. Systematic Reviews. 2014;3(1):82.

48. Guyatt G, Oxman AD, Akl EA, Kunz R, Vist G, Brozek J, et al. GRADE guidelines: 1. Introduction—GRADE evidence profiles and summary of findings tables. Journal of clinical epidemiology. 2011;64(4):383–94.

## References

1. Bolarinwa OA. Principles and methods of validity and reliability testing of questionnaires used in social and health science researches. Niger Postgrad Med J. 2015;22(4):195–201.

2. Souza AC, Alexandre NMC, Guirardello EB. Psychometric properties in instruments evaluation of reliability and validity. Epidemiol Serv Saude. 2017;26(3):649–59.

3. Bernaud J-L. Introduction à la psychométrie. Paris: France: Dunod; 2007.

4. Newton PE, Shaw SD. Validity in Educational & Psychological Assessment. London: SAGE; 2014. 280 p.

5. Hu LT, Bentler PM. Cutoff criteria for fit indexes in covariance structure analysis: Conventional criteria versus new alternatives. Structural Equation Modeling: A Multidisciplinary Journal. 1999;6(1):1–55.

6. Fokkema M, Greiff S. How Performing PCA and CFA on the Same Data Equals Trouble. European Journal of Psychological Assessment. 2017;33(6):399–402.

7. Statistics How To: Statistics for the rest of us! Kaiser-Meyer-Olkin (KMO) Test for Sampling. 2016 [Available from: https://www.statisticshowto.datasciencecentral.com/kaiser-meyer-olkin/.

8. IBM Knowledge Center. KMO and Bartlett’s Test [Available from: https://www.ibm.com/support/knowledgecenter/en/SSLVMB_24.0.0/spss/tutorials/fac_telco_kmo_01.html.

9. Edwards JR, Bagozzi RP. On the Nature and Direction of Relationships Between Constructs and Measures. Psychological Methods. 2000;5(2):155–74.

10. Matsunaga M. How to Factor-Analyze Your Data Right: Do’s, Don’ts, and How-To’s. International Journal of Psychological Research. 2010;3(1):97–110.

11. Fabrigar LR, MacCallum RC, Wegener DT, Strahan EJ. Evaluating the Use of Exploratory Factor Analysis in Psychological Research. Psychological Methods. 1999;4(3):272–99.

12. Izquierdo I, Olea J, Abad FJ. Exploratory factor analysis in validation studies: uses and recommendations. Psicothema. 2014;26(3):395–400.

13. Velicer WF. Determining the number of components from the matrix of partial correlations. Psychometrika. 1976;41(3):321–7.

14. Ellis J. Factor analysis and item analysis. Applying Statistics in Behavioural Research. Amsterdam: Boom; 2017. p. 520.

15. IBM Knowledge Center. Factor Analysis Extraction [Available from: https://www.ibm.com/support/knowledgecenter/en/SSLVMB_24.0.0/spss/base/idh_fact_ext.html.

16. Brown JD. Choosing the right type of rotation in PCA and EFA. JALT Testing & Evaluation SIG Newsletter. 2009;13(3):20–5.

17. IBM Knowledge Center. Rotation d’analyse factorielle [Available from: https://www.ibm.com/support/knowledgecenter/fr/SSLVMB_23.0.0/spss/base/idh_fact_rot.html.

18. Kenny DA. Measuring Model Fit 2015 [Available from: http://davidakenny.net/cm/fit.htm.

19. Hooper D, Coughlan J, Mullen MR. Structural equation modelling; Guidelines for determining model fit. J Res Natl Inst Stand Technol. 2008;6(1):53–60.

20. Weston R, Gore PA. A Brief Guide to Structural Equation Modeling. The Counseling Psychologist. 2016;34(5):719–51.

21. Newsom JT. Some clarification and recommendations on fit indices. 2018.

22. Podsakoff PM, MacKenzie SB, Lee JY, Podsakoff NP. Common method biases in behavioral research: a critical review of the literature and recommended remedies. J Appl Psychol. 2003;88(5):879–903.

23. Granger CV. Rasch Analysis is Important to Understand and Use for Measurement Buffalo 2008 [Available from: https://www.rasch.org/rmt/rmt213d.htm.

24. McDowell I. Measuring Health: A guide to rating scales and questionnaires Oxford University Press; 2006.

25. Reise SP, Waller NG, Comrey AL. Factor analysis and scale revision. Psychological assessment. 2000;12(3):287.

26. Cameron AC, Trivedi PK. Microeconometrics: Methods and applications. New York: Cambridge University Press; 2005. 1034 p.

27. Langevin V, François M, Boini S, Riou A. Les questionnaires dans la démarche de prévention du stress au travail.. Documents pour le Médecin du Travail. 2011;125(1er trimestre 2011):23–35.

28. NCSS Statistical Software. Canonical correlation. Unkown [Available from: https://ncss-wpengine.netdna-ssl.com/wp-content/themes/ncss/pdf/Procedures/NCSS/Canonical_Correlation.pdf.

29. Henseler J, Ringle CM, Sarstedt M. A new criterion for assessing discriminant validity in variance-based structural equation modeling. Journal of the Academy of Marketing Science. 2014;43(1):115–35.

30. Liu L, Li C, Zhu D. A new approach to testing nomological validity and its application to a second-order measurement model of trust. Journal of the Association for Information Systems. 2012;13(12):950–75.

31. Warrens M. On Cronbach’s Alpha as the Mean of All Split-Half Reliabilities. Quantitative Psychology Research. 89: Springer Proceedings in Mathematics & Statistics; 2015.

32. Zumbo BD, Gadermann AM, Zeisser C. Ordinal Versions of Coefficients Alpha and Theta for Likert Rating Scales. Journal of Modern Applied Statistical Methods. 2007;6(1):21–9.

33. Statistics How To: Statistics for the rest of us! Guttman’s lambda-2: Definition, Examples [Available from: https://www.statisticshowto.datasciencecentral.com/guttmans-lambda-2/.

34. Michalos AC. Encyclopedia of Quality of Life and Well-Being Research. Prince George, BC, Canada: SpringerReference; 2014. 7347 p.

35. Koo TK, Li MY. A Guideline of Selecting and Reporting Intraclass Correlation Coefficients for Reliability Research. J Chiropr Med. 2016;15(2):155–63.

36. Mandrekar JN. Measures of interrater agreement. Journal of Thoracic Oncology. 2011;6(1):6–7.

37. Statistics How To: Statistics for the rest of us! Standard Error of Measurement (SEm): Definition, Meaning. 2016 [Available from: https://www.statisticshowto.datasciencecentral.com/standard-error-of-measurement/.

38. Vacha-Haase T, Thompson B. How to Estimate and Interpret Various Effect Sizes. Journal of Counseling Psychology. 2004;51(4):473–81.

39. MEDALC easy-to-use statistical software. Responsiveness. Unkown [Available from: https://www.medcalc.org/manual/responsiveness.php.

40. Becker LA. Effect Size (ES). 2000.

